# Beyond prescribed activities: examining passive postural sway and gait data in patients with multiple sclerosis

**DOI:** 10.1101/2024.11.05.24316692

**Authors:** Brett M. Meyer, Nishit Agarwal, Kevin Machado Gamboa, Aisling O’Learly, Andrew J. Solomon, Ryan S. McGinnis, Melissa Ceruolo

## Abstract

Symptoms of multiple sclerosis (MS) are highly variable and include impaired senses, instability, and fatigue, making persons with MS (PwMS) ill-suited for the traditional six-month office visit paradigm. Instead, PwMS are well suited for remote monitoring to capture their true impairment. The objective of this work is to investigate the value of free-living data compared to prescribed walking tasks. Wearable sensor data were utilized from six-weeks of data from 25 PwMS. Participants completed a daily 1-minute walk, 30-second standing task, and patient reported outcomes of balance confidence (ABC), fatigue (MFIS) and walking impairment (MSWS). We compared gait and sway, as well as correlated them to patient reported outcomes (PRO). Lastly, we used a regression to determine the variance accounted for (VAF) in each PRO by different data sources. Temporal gait features were moderately correlated (r=0.6 – 0.81) between passive and prescribed walking, however, no postural sway features were correlated with each other. Passive data was found to have greater clinical relevance in our sample of PwMS compared to prescribe tasks for both gait and sway analyses. Passive sway features were found to be moderately related to ABC, MFIS, and MSWS (r=0.42-0.74, VAF=0.42-0.7), while prescribe sway was only correlated to MFIS (r = 0.41, VAF = 0.44). Both passive and prescribed measures of gait were related to ABC and MSWS; stronger relationships were found in the passive data (r = 0.42-0.78, VAF = 0.64-0.78). Additionally, we found the performance increased for passive monitoring with a shorter monitoring duration – highlighting the need to properly match the monitoring and analysis duration to the population. Overall, our findings highlight the importance of including passive free-living analysis in future studies.

## Introduction

Multiple Sclerosis (MS) is a chronic disease of the central nervous system characterized by progressive demyelination and axonal damage (1). People with MS (PwMS) often experience symptoms such as fatigue, impaired sensation, and reduced muscle strength and coordination (2). These symptoms can lead to difficulties with postural control and dynamic activities like standing and walking. Among the estimated 2.8 million PwMS worldwide, 50-80% report issues with gait and balance, with over half experiencing a fall each year (1,3,4). Notably, these impairments can fluctuate over time, varying from hours to days or weeks. This variability makes it challenging to prescribe effective interventions, such as walking aids, using the traditional care model of six-month office visits (5). As a result, PwMS are well-suited for continuous free-living monitoring to better assess balance and gait impairments.

Advances in technology now enable free-living assessments through various methods. These range from smartphone applications that track activity and health status, to camera-based systems for monitoring specific activities, to wearable inertial measurement units (IMUs) that capture body movement (6). Each technology has its trade-offs, but for this study, we focus on wearable IMUs, which offer a cost-effective way to measure movement while posing fewer privacy concerns than camera-based systems(7,8). Despite their increasing use, few studies have examined the best methods for utilizing wearables in long-term monitoring. Research has shown that walking in everyday life differs significantly from walking in a clinical setting, and the required duration of monitoring depends on variability and frequency of activity (9,10). However, it remains unclear whether prescribed activities, such as walking or balance assessments performed remotely, are sufficient, or if passive data collection during daily activities is also necessary.

Previous research has shown that both prescribed and free-living assessments of walking and balance yield clinically relevant outcomes. In-clinic walking tests have been strongly linked to fall risk and are more effective at classifying fall risk than free-living walking with current analysis techniques (10,11). While free-living walking is associated with clinical outcomes and fall status, the accuracy of fall risk classification depends on the length of the walking period analyzed (10). In-clinic balance assessments, often referred to as postural sway tests, are commonly used and have been shown to correlate strongly with fall risk and disease severity across various populations (12–14). Free-living assessments of postural sway are an emerging field and have demonstrated clinical significance, but these relationships depend on the context in which the standing data is collected. In addition, several studies have shown that many prescribed tasks, such as walking and functional assessments, can be performed at home (15,16).

Given the variability in MS symptoms, continuous monitoring offers clear benefits for PwMS, but there is a need to learn how this can effectively be implemented in clinical care or research. The objective of this study is to compare the clinical significance of gait and balance measures between prescribed remote tasks and free-living daily life data. We aim to provide recommendations for incorporating free-living monitoring into wearable sensor studies.

## Materials and Methods

### Dataset: Subjects and Protocol

The study presented herein is a secondary analysis of previously collected data (17,18). During data collection, we recruited 25 individuals with multiple sclerosis (MS) (5 males, 20 females; mean age 50 ± 9.7 years) from the University of Vermont Medical Center’s MS Center and the IDEAL for MS Program. Participants were included if they had no conditions other than MS affecting balance or mobility, were able to walk unaided, were not pregnant or breastfeeding, and had no known skin sensitivities to adhesives or hydrogel.

Participants were monitored biweekly for 12 weeks, generating six weeks of sensor data for analysis (Fig 1.). All participants completed at least two weeks of monitoring (n=25), with 23 completing five weeks and 22 completing the full six weeks. During the sensor weeks, participants performed a daily 30-second chair stand test, a one-minute walk, and a 30-second standing balance assessment. During the one-minute walk, participants were instructed to walk continuously for 1 minute at a comfortable pace for the prescribed walk. During the prescribed standing trial, participants were instructed to stand upright and still for 30 seconds with their feet shoulder-width apart and eyes open. In addition, they filled out daily falls surveys and the Activity-Specific Balance Confidence (ABC) questionnaire. On non-sensor weeks, participants completed the Modified Fatigue Impact Scale (MFIS) and the 12-item Multiple Sclerosis Walking Scale (MSWS), which aligned with the recall periods of these assessments.

**Fig 1.**
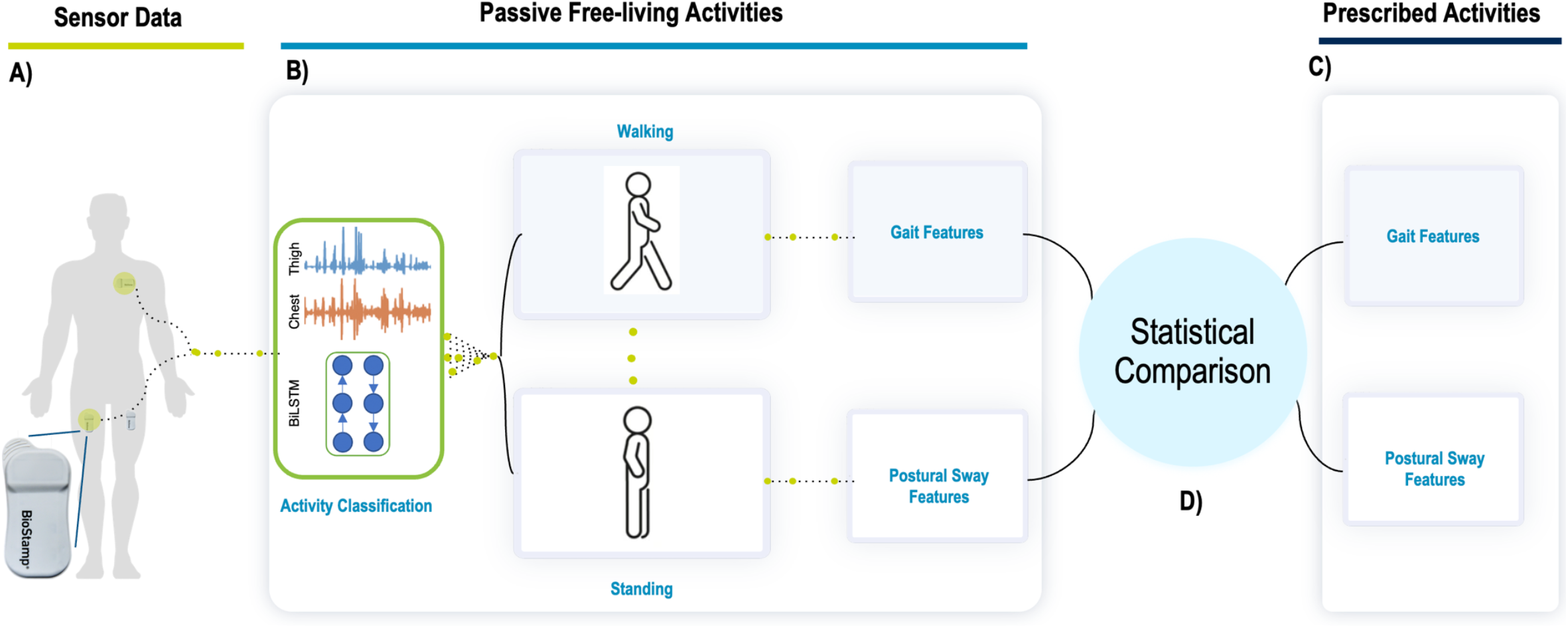
Overview of study analysis. A) Data was collected from BioStamp nPoint (Medidata) wearable sensors located on the thigh and chest continuously for up to six weeks. B) An activity classification model was used to identify walking and standing periods from which features of gait and postural sway were extracted. C) Participants were asked to complete a daily one-minute walk and 30-second standing period. Features of gait and postural sway were extracted from these prescribed activities. D) Features derived from the passive free-living activities were compared the features derived from prescribed activities using difference tests and relationships to patient reported outcomes.

Participants wore BioStamp nPoint sensors during sensor weeks, placed on the left upper chest and both thighs. The sensors recorded acceleration (31.25 Hz, ±16G) and electromyography (250 Hz) data continuously throughout the day. Data was stored locally on the sensors and uploaded to the cloud daily via a docking station provided to each participant. At the conclusion of the study, participants completed the Patient-Determined Disease Steps (PDDS) scale to assess disability levels. Survey results for the cohort were as follows: PDDS 0.88 (1.05); ABC 77.6 (21.9); MFIS 28.3 (16.1); MSWS 19.1 (7.1). This study protocol received approval from the University of Vermont’s Institutional Review Board (CHRMS 21-0401).

### Activity Classification

Free-living walking and standing periods were identified from the wearable accelerometer data using a previously validated classification model (13,16,17). This deep learning model uses raw accelerometer data from sensors placed on the thighs and chest to classify activities into categories such as walking, standing, sitting, lying, or other. The model employs two Long-Short-Term-Memory (LSTM) layers and was trained on over 100,000 four-second segments of data from individuals with MS, healthy adults, and those with Parkinson’s disease, achieving a validation accuracy of over 96% (16).

### Gait Feature Extraction

Once activity classification was complete, gait events were identified in walking bouts using a thigh-based acceleration method detailed in previous studies (19,20). Walking bouts containing at least two valid strides were analyzed. Several temporal, asymmetry, and stability measures were extracted from each walking bout. Temporal gait parameters, calculated for each stride and averaged across the bout, included stride duration, stance duration, swing duration, duty factor, and double support duration (20).

Asymmetry measures included duty factor asymmetry (normalized using the L1-norm, 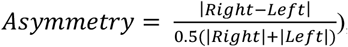, correlation asymmetry (an affine transformation of the correlation between right and left thigh acceleration), and acceleration asymmetry (calculated using an L1-norm method between ensemble averages of right and left stride acceleration) (20). Stability measures included the root-mean-square (RMS) of anterior-posterior (AP) acceleration, frequency dispersion of medial-lateral (ML) acceleration (21), entropy ratio between the trunk and thigh (22), and the Lyapunov exponent in both AP and ML directions (21). Entropy ratio was only calculated for walking bouts longer than 30 seconds, while the Lyapunov exponent was calculated for bouts lasting more than one minute. Entropy ratio asymmetry was also calculated using the L1-norm method. The selected features are supported by existing literature, which has linked them to MS-related gait impairment and fall risk (10,17,21–23).

### Postural Sway Feature Extraction

Postural sway features were extracted from standing periods lasting at least 30 seconds, following established methods(12,24). These standing bouts were used to compute sway-related parameters, including sway area (normalized by bout duration), centroidal frequency, sway distance, and key frequency characteristics (e.g., 50th and 95th percentiles of frequency content, frequency dispersion). Additional measures such as jerk, mean period, mean velocity, normalized path, power in the frequency spectrum, range of acceleration, and RMS acceleration were also calculated. Sway distance thresholds were applied to remove standing bouts where participants were completely still or exhibited erratic movements. These thresholds were based on the 1.5th and 98.5th percentiles of 10,000 randomly selected unfiltered standing bouts and confirmed with manual inspection, ensuring only valid standing periods were analyzed.

All gait and sway features were computed for passive free-living data and the prescribed tasks using custom MATLAB scripts executed in Medidata’s SensorCloud production environment.

## Statistical Analysis

Gait and sway features were analyzed for the full six-week study period as well as a shorter three-day monitoring period, as recommended by prior studies (17). Gait analysis was further separated into two free-living groups, one containing all free-living (all-free) passive gait and one containing only gait features from passive walking periods that were 30 seconds or longer (long-free) (10). For each participant, features were aggregated using the median, 25th percentile, and 75th percentile for both prescribed and passive data sources. Statistical comparisons were performed using the Mann-Whitney U test to assess differences between conditions, while Spearman correlation was used to evaluate associations between features from different activity domains. Comparisons were made between passive and prescribed walking features, and between standing features during passive and prescribed activities.

To assess the clinical relevance of these features, Spearman correlations were computed between the extracted features and patient-reported outcomes for balance confidence (ABC), fatigue (MFIS), and walking impairment (MSWS) using the median of each PRO. The overall significance of features from each domain was evaluated by measuring the variance accounted for (VAF). Principal component analysis (PCA) was used to reduce the feature set, retaining the components necessary to explain 95% of the variance. For all passive and long passive gait features, seven principal components were required, while for prescribed walking, five components were needed for both the full six-week and three-day monitoring periods. Similarly, for prescribed standing, seven principal components were required for both monitoring durations, while five components were necessary for passive standing over six weeks and seven components for the three-day period.

## Results

Comparing the results of difference tests and clinical relevance between the different data sources, passive free-living gait and sway are significantly different from the gait and sway collected from prescribed walking and standing tasks. Out of the 42 feature and aggregation combinations, difference tests of gait find six significant differences between long-free living and prescribed walking. When considering all free-living walking, this difference increases to 26 significantly different features. Reducing the study duration to three days, we find similar results of five and 25 significantly different features between long-free and all free-living walking to prescribed walking, respectively. The features and aggregations most similar are depicted in Fig 2. Further analysis of Fig 2 reveals that weak to moderate correlations exist between gait timing, asymmetry, and stability features when considering the full study duration, however, many of these correlations weaken when considering three days of data.

**Fig 2.**
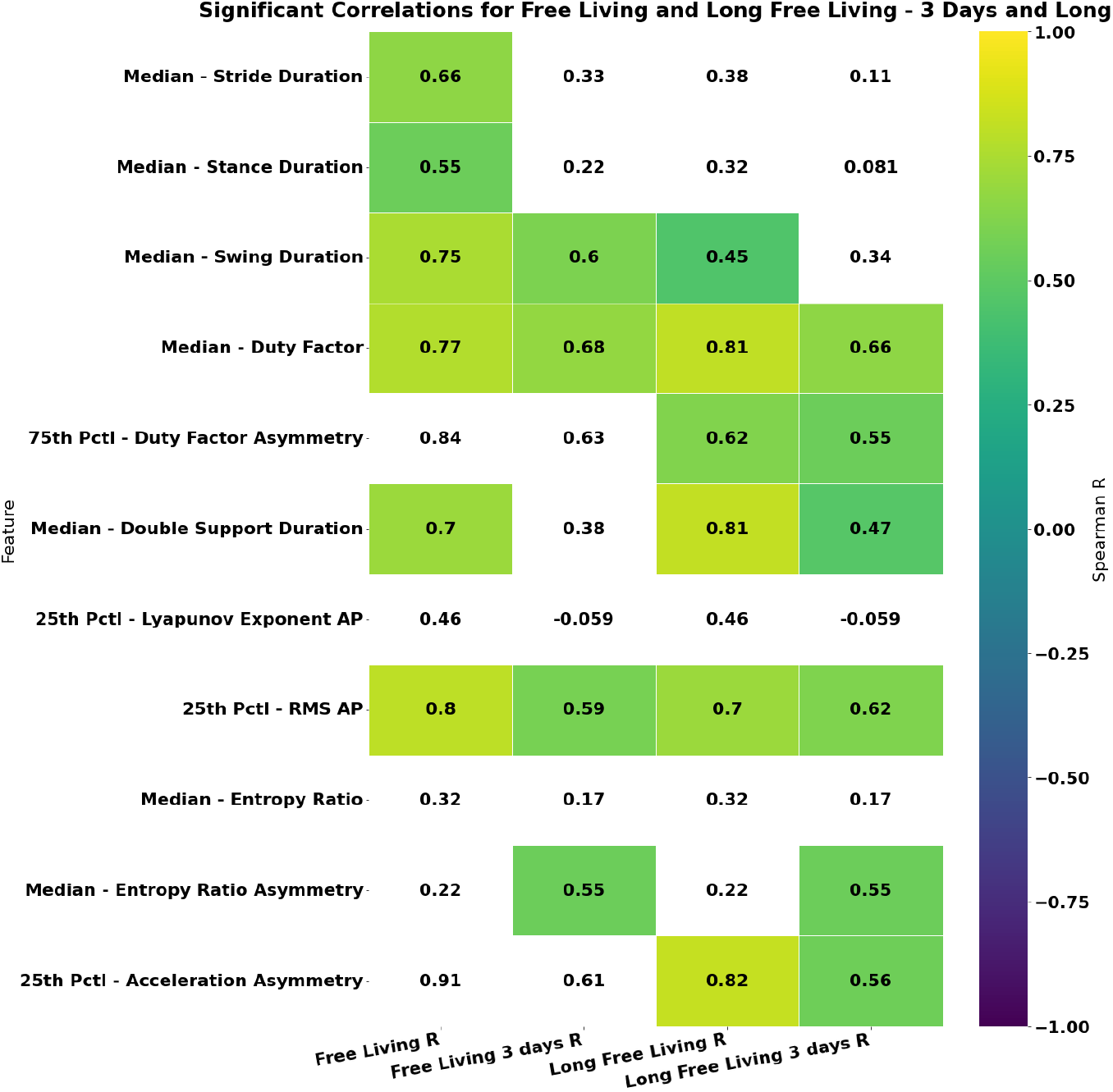
Comparison of strongest relationships found between gait features from prescribed walking and passive free-living walking gait features. Long signifies only gait bouts over 30 seconds are considered. Tests performed considering full six-week study and shorter three-day period. Spearman correlation displayed. Features with non-significant correlations or significant differences in medians are displayed as white. Each column represents the correlation to the prescribed feature from either all free-living gait or long free-living gait from either the full six-week study or three-day period.

Comparing the clinical relevance of the different data sources of gait, passive monitoring establishes stronger correlations overall to the patient reported outcomes (PROs) as seen in Fig 3. First looking at ABC, we find far more and stronger significant correlations between balance confidence and gait from free-living data compared to prescribed walking (Fig 3A). The shorter three-day duration highlights this further, where nearly all relationships that existed between prescribed gait and ABC disappear, meanwhile these relationships are retained in free living data. In this case all free-living walking also outperformed the reduced set of longer walks on both study durations.

**Fig 3.**
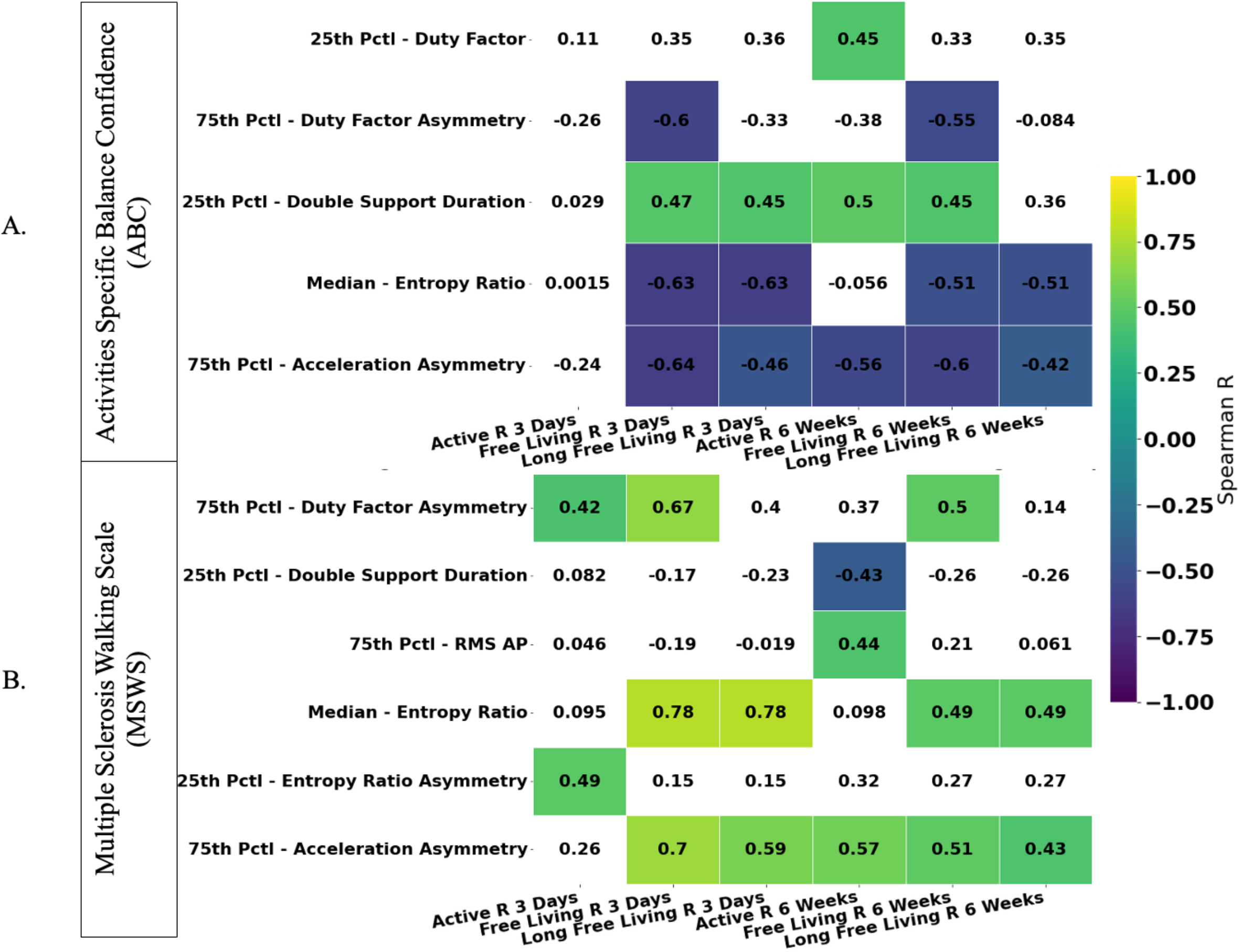
Spearman correlation between patient reported outcomes and gait features derived from prescribed (active), long free living bouts, and all passive free-living bouts of walking data. Reduced feature set displayed by selecting the strongest correlation per feature aggregation. Non-significant correlations displayed in white. Tests performed using full six-week study data and reduced three-day monitoring window. Each column represents the correlation to the prescribed feature from passive free-living sway from either the full six-week study or three-day period.

When considering MSWS from the full study duration, we find the strongest correlation with acceleration asymmetry found from prescribed walking (r=0.57), however, on average free-living correlations are stronger (Fig 3B). Reducing to three days of data, we saw much stronger correlations between free-living gait than observed from the passive walks in both study durations. The correlation to acceleration asymmetry increases to 0.7 (from 0.57) and the strongest correlation was 0.78. Interestingly, the strongest and only significant relationship between MFIS was also found with three-days of the passive free-living gait (Entropy Ratio, r = 0.5).

Unlike gait, all sway features were found to be significantly different between the passive free-living standing and the prescribed standing task. These data are very different from each other and have very different clinical relevance. All significant correlations to ABC were found with the free-living data when considering both study durations as seen in Fig 4A. The strongest relationships were found between ABC and sway range and distance when considering three-days of data; the strength of these correlations is reduced for the full study duration. The same results are seen between sway features and MSWS; no significant correlations with the prescribed standing and the strongest correlations are found from three-days of passive free-living standing as seen in Fig 4B. The strongest observed correlation was the 75^th^ percentile of sway range (r = 0.7). When considering MFIS, passive sway range continues to provide the strongest correlations (Fig 4C) and passive data provides more and stronger significant correlations, however, there is a unique relationship to the 50^th^ Percentile Frequency found in the prescribed standing task.

**Fig 4.**
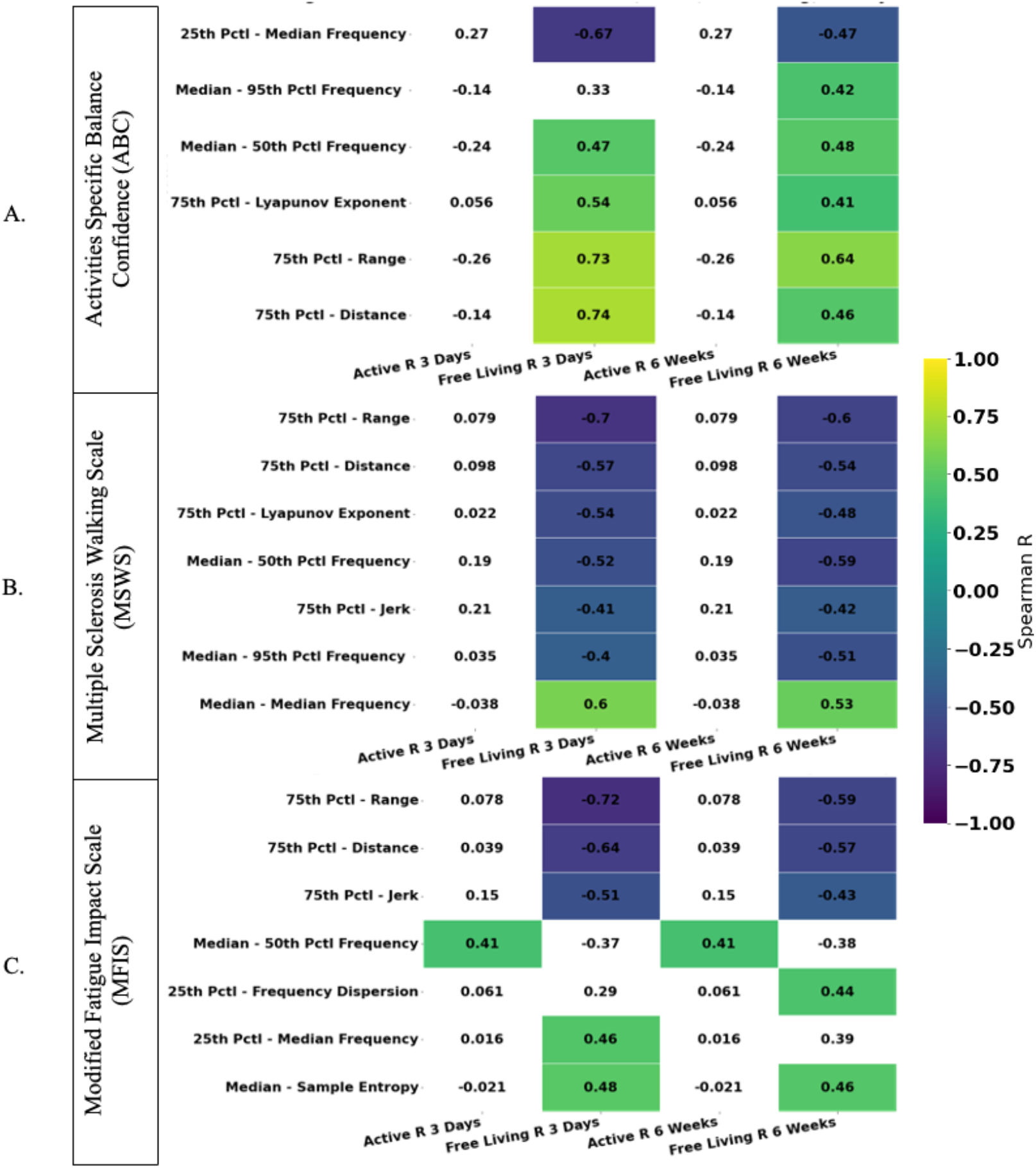
Spearman correlation between features of postural sway from prescribed (active) and passive free-living standing data to patient reported outcomes. Reduced feature set displayed by selecting the strongest correlation per feature aggregation. Non-significant correlations displayed in white. Tests performed using full six-week study data and reduced three-day monitoring window.

Moving beyond individual feature correlations, we also used PCA and performed a regression for each data source to determine the amount of variance explained as seen in Table 1. As found in the individual feature analysis, we find the most variance in the PROs is explained by passive walking and standing collected over three-days. All passive free-living walking explains the most variance in ABC and MSWS, 0.76 and 0.64, respectively, while passive standing explains the most variance in MFIS, 0.61. Utilizing the full study duration reduces variance explained for each data source, however, passive data is still found to outperform both prescribed walking and standing.

**Table 1.** Variance Accounted for in Patient Reported Outcomes.

| Patient Outcome | Prescribed Walking |  | Long Passive Walking |  | All Passive Walking |  | Prescribed Standing |  | Passive Standing |  |
| --- | --- | --- | --- | --- | --- | --- | --- | --- | --- | --- |
|  | 3 Days | 6 Weeks | 3 Days | 6 Weeks | 3 Days | 6 Weeks | 3 Days | 6 Weeks | 3 Days | 6 Weeks |
| ABC | 0.42 | 0.57 | 0.71 | 0.61 | **0.76** | 0.64 | 0.16 | 0.16 | **0.70** | 0.36 |
| MSWS | 0.15 | 0.35 | 0.58 | 0.38 | **0.64** | 0.38 | 0.09 | 0.09 | **0.42** | 0.36 |
| MFIS | 0.15 | 0.08 | 0.25 | 0.15 | **0.49** | 0.12 | 0.43 | 0.44 | **0.61** | 0.47 |

Variance accounted for in a regression model predicting patient reported outcomes from postural sway and gait features. Principal component used to reduce each feature set. Number of components required to explain 95% of variance were used in the regression. Tests performed using full six-week study data and reduced three-day monitoring window. ABC: Activities Specific Balance Confidence; MSWS: Multiple Sclerosis Walking Scale; MFIS: Modified Fatigue Impact Scale

## Discussion

The purpose of this study was to investigate the benefit of passive free-living data analysis and compare its value to free-living prescribed tasks of walking and standing. In doing so, we first compared the data derived from the passive and prescribed activities. We then computed correlation to patient reported outcomes to establish the relevance of the individual features from each data source. Lastly, we used a regression approach to determine the relevance of the features as a group and computed the variance-accounted-for from each data source. Consistently throughout these analyses, we saw prescribed and passive free-living data are different from each other and have very different relationships to clinical measures in both gait and sway analyses.

Based on previous studies, we split our passive gait analyses into two groups; long passive walking bouts (over 30 seconds) and all passive walking bouts (10). As expected, the long passive walking bouts were more similar to the prescribed walking than all passive walking and worsening impairment was associated with worsening stability and asymmetry features. Unlike previous studies that found longer walking is more predictive of fall risk, our analyses found greater clinical revelance across all correlations and variances explained when considering all-passive data compared to long passive and prescribed walking for each PRO (10). We also consistently found that considering three days of data instead of up to six weeks provided more clinically significant data in this population.

Our analysis of passive and prescribed standing yielded the same results, however, the discrepancies between the data sources was even greater. In most instances, the prescribed standing task demonstrated little clinical relevance, where the passive standing analysis demonstrated strong correlations and explained far more variance. The features Range and Distance provided strong correlations to each PRO. Interestingly, higher sway range and distance was indicative of less impairment in each PRO. This is consistent with previous findings and suggests the passive standing reveals less impaired PwMS may perform more dynamic activities while standing at home (13). We hypothesize that these dynamic activities may be responsible for this large difference between passive and prescribed sway and highlight disease characteristics more than a simple standings task. Additionally, due to the remote nature, we cannot rule out improper task performance such as holding onto or leaning on something. Like our findings in gait, the shorter three-day monitoring duration was also found to yield more clinically relevant relationships than the full six-week study.

As demonstrated consistently across gait and sway, considering three-days of passive data has greater clinical relevance than considering the full six-weeks of data. The opposite was found for prescribed gait, where the relationships that had developed over the full study were not formed in three days. We hypothesis that this is because three days of one walking bout each are not enough to characterize the gait of PwMS. On the contrary, six-weeks may be too much passive data to consider for PwMS given their symptom fluctuations (25). This finding highlights the importance of matching appropriate study design and analyzes with the disease being studied.

In this analysis we demonstrated that passive free-living measures of gait and sway and highly related to patient reported measures of balance confidence, fatigue, and walking impairment. These objective measures of movement are related to established clinically relevant patient reported outcomes. This satisfies final step of the V3 method of determining if digital measures are fit for purpose, clinical validation (26), furthering the evidence of the clinical significance of free-living analysis.

There are some limitations to this current study. Our sample size of 25 participants limits our statistical power and due to COVID-19, participants were not able to be assessed by a neurologist to obtain more in-depth disease characteristics. Due to our inclusion criteria of being able to walk unaided, we also have a lower impairment sample of PwMS. Further studies will be needed to confirm this in PwMS with great disease burden. Additionally, since we found stronger results from a shorter monitoring duration, longitudinal analysis of passive data from PwMS may require repeated measures analysis. Lastly, our prescribed tasks instructed patients to walk and stand comfortably for the duration of the task, therefore, future studies will be needed to confirm whether these findings are consistent with more challenging activities like tandem standing or a six-minute walk test. Despite these limitations, our findings clearly demonstrate the importance of passive free-living monitoring in study design. When considering data from both gait and postural sway, passively collecting data from people’s daily lives more closely aligns with patient reported outcomes than asking them to perform a prescribed task.

## Conclusion

Here we investigated the importance of passive free-living data collection and analysis in PwMS. We compared walking and standing data collected during six-weeks of sensor wear derived from prescribed daily standing and walking tasks to passive data from participants’ daily activities. Passive data was found to have greater clinical relevance in our sample of PwMS compared to prescribe tasks for both gait and sway analyses. Additionally, we found the performance increased with a shorter monitoring duration – highlighting the need to properly match the monitoring and analysis duration to the population. Overall, our findings highlight the importance of including passive free-living analysis in future studies.

## Data Availability

Data is currently unavailable

